# Longitudinal changes in processed foods intake and their daily caloric contribution among Ghanaian populations living in Ghana and Europe: Findings from the prospective RODAM Cohort study

**DOI:** 10.1101/2025.06.06.25329107

**Authors:** Mustapha Titi Yussif, Reginald Adjetey Annan, Anthony Edusei, Mary Nicolaou, Erik Beune, Samuel Nkansah Darko, Ina Danquah, Karlijn A.C. Meeks, Ellis Owusu-Dabo, Charles Agyemang

**Affiliations:** Department of Biochemistry and Biotechnology, Faculty of Biosciences, College of Science, Kwame Nkrumah University of Science and Technology, Kumasi, Ghana; Department of Public and Occupational Health, Amsterdam Public Health Research institute, Amsterdam University Medical Center, University of Amsterdam, Amsterdam, The Netherlands; School of Public Health, Kwame Nkrumah University of Science and Technology (KNUST), Kumasi, Ghana; Heidelberg Institute of Global Health (HIGH), Universitätsklinikum Heidelberg, Medizinische Fakultät der Universität Heidelberg, Heidelberg, Deutschland; Hertz-Chair Innovation for Planetary Health, Zentrum für Entwicklungsforschung (ZEF), Universität in Bonn, Bonn, Deutschland; Division of Endocrinology, Diabetes and Nutrition, Department of Medicine, University of Maryland School of Medicine, Baltimore, MD, USA; Division of Endocrinology, Diabetes, and Metabolism, Department of Medicine, Johns Hopkins University School of Medicine, Baltimore, MD, USA; Department of Molecular Medicine, Kwame Nkrumah University of Science and Technology (KNUST), Kumasi, Ghana

**Keywords:** processed foods, ultra-processed foods, energy, NOVA, Ghana

## Abstract

**Background:** Dietary changes towards an increased consumption and caloric contribution of processed and ultra-processed foods are major causes of obesity and non-communicable diseases worldwide. However, data on the intake of processed foods among Ghanaians living in Ghana and their migrant counterparts living in Europe, which could help assess the impact of urbanization and migration on diet and health outcomes, are limited. Therefore, the present study assessed changes in the intake of processed foods and their corresponding energy contribution among different Ghanaian populations.

**Methods:** Dietary data were collected on Ghanaians from the RODAM-Pros (Research on Obesity and Diabetes among African Migrants - prospective) cohort study, which recruited Ghanaians living in rural and urban Ghana and also Ghanaian migrants living in Amsterdam between 2011-2015 and 2019-2021. Dietary intake data were analyzed by regrouping foods based on the extent and purpose of processing according to the NOVA classification. The paired sample t-tests were conducted to determine the differences in mean daily intake (grams) of foods and energy between baseline and follow-up. Energy values of the various foods were retrieved from West African food composition data.

**Results:** Compared with the baseline data, a significant increase in the consumption of ultra-processed foods was found among migrant Ghanaians living in Amsterdam (72.7% increase, Mean Diff: 154.97g/day, 95%CI: 115.39 −194.54, p<0.001) with no significant changes among their rural and urban counterparts. The change in percentage of total energy from ultra-processed foods was 9.6% to 9.0% (p= 0.136), 15.9% to 13.9% (p<0.001) and 13.4% to 13.0% (p= 0.539), respectively, among rural, urban Ghanaians and Ghanaian migrants in Amsterdam.

Processed food consumption on the other hand increased across all study sites by 53.4% (Mean Diff: 151.12g/day, 95%CI: 129.56 −172.68, p<0.001) in rural Ghana, 33.1% (Mean Diff: 100.62g/day, 95%CI: 79.08-122.16, p<0.001) in urban Ghana and by 220.0% (Mean Diff: 187.66g/day, 95%CI: 174.72 −200.59, p<0.001) among migrants living in Amsterdam, the Netherlands. Similarly, the percentage contribution from processed foods to total energy increased significantly from baseline data to endpoint among all population groups, thus, 20.8% to 39.5% (p<0.001) among rural dwellers, 23.6 % to 33.5% (p<0.001) among urban dwellers and 7.2 % to 12.9% (p<0.001) among migrant Ghanaians in Amsterdam.

**Conclusion:** Dietary intake has shifted towards higher intake of ultra-processed and processed foods with caloric contribution from processed foods having increased for all Ghanaian population groups. The importance of the observed changes in the consumption of processed and ultra-processed food for the risk of cardio-metabolic diseases among Ghanaian populations remains to be evaluated

## INTRODUCTION

Processed and ultra-processed foods are mainly characterized by high energy and low micronutrients density [1,2] as such dietary habits that favour processed foods consumption pose a high threat to human health [3]. Evidence abounds that ultra-processed foods are linked to Non-Communicable Diseases (NCDs) such as the obesity pandemic [4], cancer [5], cardiometabolic disorders [6,7], and mortality [8].

In Sub Saharan African (SSA) Countries, there has been a rise in NCDs, which is believed to be driven in part by the nutrition transition where traditional diets are giving way to processed and ultra-processed food consumption which has been predicted to continue to rise in these countries if no policy interventions are implemented [9,10].

Ghana is one of the countries in the sub-Saharan Africa where changes in dietary intake and habits have been noticed since the early 1990s [11]. The country has been suggested to be in the later stage of the nutrition transition with its attendant repercussions on health [12]. Recent epidemiological studies have shown high levels of obesity, hypertension and other related cardiovascular diseases (CVDs) among Ghanaians with significant variations among population sub-groups based on migration and geographical location [13–15] making the investigation of the role of processed foods in the nutrition transition worthy.

It is yet unclear what changes in dietary intake could have accounted for these variations in CVD outcomes in these population sub-groups underpinning the need to consider contextual differences in food consumption.

In Ghana, the use of processed foods may be increasing under the influence of an obesogenic food environment [16,17]. Even in rural areas, access to and consumption of processed and ultra-processed food may be rising due to a number of factors, including increased availability and changing lifestyles [18], greater incomes from non-farm jobs, mechanization of farm output, and an increase in demand for convenience [19]. There is also increasing urbanization, which could lead to increased consumption of processed foods as a result of the link between urbanization and higher incomes and employment opportunities. The growing time demands and the rise in the demand for convenience foods, which may be caused among other things by changes in the workforce and family patterns, notably the increasing economic participation of women, which leads to a rise in demand for convenience foods [20].

In the advanced economies, findings from several observational studies reported a high intake of dietary energy from processed foods compared with unprocessed foods. For instance, in the United States, a previous study found that ultra-processed foods accounted close to 58% of dietary energy among adults [21]. Similarly, ultra-processed foods contributed 55% of calories among Canadians [22]. Further, in the United Kingdom, a study revealed that 51% of calories were from ultra-processed foods [23]. Moreso, a large study in 10 European countries revealed that processed foods contributed 61-79% of energy intake in Spain, the Netherlands and Germany [24]. This suggests that Ghanaian migrants in Europe and other western countries may increase their consumption of processed foods due to dietary acculturation where they adopt to diets that are higher in processed foods. However, little is known about the quantity of processed foods intake and the corresponding dietary energy intake from these processed foods and how these are changing over time. If the levels of dietary intake of processed foods and its trend among rural, urban and migrant Ghanaian groups are known then we can assess how the availability, affordability and marketing of processed foods differ within the different Ghanaian contexts. It will also help to assess the direct impact of urbanization and migration on diet and health outcomes. It would be known whether migrants retain their traditional diets or adopt host-country food patterns, and how these choices affect their health. Understanding these dietary trends can guide health interventions, policies, and educational programs to promote healthier eating habits among these populations.

Therefore, the present study assessed the changes in intake of ultra-processed foods, processed foods, and unprocessed/minimally processed foods, and their corresponding daily caloric contribution across three different Ghanaian population groups living in rural and urban Ghana, and their migrant counterparts living in Amsterdam, the Netherlands over a six-year period.

## METHODS

### Data Source and Collection

In this study we used data from the Research on Obesity and Diabetes among African Migrants (RODAM) prospective cohort study (RODAM-Pros) of which the details related to the participant recruitment process and data collection techniques have been published elsewhere [25,26]. In brief, the RODAM-Pros cohort study was conducted to assess the drivers of the high burden of CVD risk by assessing the key changes in environmental exposures and epigenetic modifications among sub-Saharan African migrants. It is a longitudinal cohort study, which collected follow-up data between 2019-2021 from participants who completed the RODAM baseline study between 2011-2015 and were eligible for follow-up.

The RODAM study baseline was based on a well-defined homogenous SSA population (i.e., Ghanaian migrants of mostly Akan ancestral heritage) living in three European cities (Amsterdam, the Netherlands; Berlin, Germany and London, UK) and their compatriots living in rural and urban Ashanti region of Ghana. The RODAM-Pros cohort was restricted to the Netherlands, rural and urban Ghana because the recruitment strategies in these sites allowed the study participants to be followed over time.

Two cities in Ghana (Kumasi and Obuasi) served as the sites for the urban areas whilst 15 villages in the Ashanti region served as the site for the rural participants. Multistage sampling procedure was used to arrive at 15 enumeration areas (EAs) for the rural sites and 15 EAs for the urban sites. Participants of Ghanaian migrants were randomly recruited for the Amsterdam study site from the Amsterdam Municipal register [25].

### Dietary Assessment and Classification of food groups

Dietary assessment was conducted using the 134-item Ghana Food Propensity Questionnaire (Ghana-FPQ) which queried for usual intake of foods in the past 12 months. The Ghana-FPQ is a standardized Questionnaire developed based on the European Food Propensity Questionnaire (EFPQ) [27] by the incorporation of typical Ghanaian foods identified from the 2008 Ghana Sociodemographic and Health Survey [28] as well as from previous studies conducted among urban dwelling Ghanaians and Ghanaians living in Amsterdam [29].

Usual daily intake of foods in grams per day was estimated by combining the frequency of intake with standard portion sizes, which was determined through the conduct of 24-hour dietary recalls among a random sub-sample of 251 participants for the Ghanaian food items and the EFPQ was used for the European foods [30].

We mapped and re-classified 30 food groups into four groups according to the NOVA international classification system based on the extent and purpose of processing [31]. The four groups were: NOVA Group 1 - unprocessed/minimally processed foods; NOVA Group 2 - Processed culinary ingredients; NOVA Group 3 - Processed foods and NOVA Group 4 – Ultra-processed foods. The full list and constitution of the food groups extracted from the Ghana-FPQ and their respective NOVA classification is attached as Supplementary Table 1.

The usual daily intakes in grams per day of the various NOVA classification groups were obtained by summing the intakes per day of food items within the respective food groups that constituted the NOVA group. Subsequently, the usual daily intakes (g/day) from each of the four NOVA groups were then translated into energy and nutrient intakes using the West African Food Composition Table (2012) [32] and the German Nutrient Database (BLS 3.01) (2010).

### Socio-demographic variables

Socio-demographic variables include sex, relationship status (married or registered partnership, cohabitating or living together, unmarried or never married, divorced or separated, widow/widower), educational status (categorized into four; never been to school or elementary school, lower vocational schooling or lower secondary schooling, intermediate vocational schooling or intermediate/higher secondary schooling, and higher vocational schooling or university), employment status (re-coded into two categories as employed and unemployed).

### Data Analysis

Baseline data were summarized as means with standard deviations for continuous variables while categorical data was summarized using frequencies and their relative percentages. The normality of the absolute food intake (grams/day) and energy (kcals/day) were assessed using the Kolmogorov-Smirnov test. These variables showed a normal distribution (Kolmogorov-Smirnov test has a statistical significance > 0.050). We compared the mean daily intake of foods (grams/day) between baseline and follow-up using paired sample t-tests. The energy contribution relative to total energy of each of the four food groups (unprocessed, culinary ingredients, processed, and ultra-processed foods) was analyzed stratified by location of residence of the three Ghanaian populations. The percentage of energy consumed from each food processing group was computed as the ratio of the energy intake from that food group to the total daily energy intake, and expressed as a percentage [33]. The differences in the percentage of calories consumed from the various food groups between baseline and follow-up were also computed using the paired samples t-test. All statistical analyses were conducted using the statistical software IBM SPSS Statistics for Windows version 24.0 and the level of significance adopted was α ≤ 0.050.

### Ethics, consent and permissions

Ethical approval was granted from the respective ethics committees in Ghana (School of Medical Sciences/Komfo Anokye Teaching Hospital Committee on Human Research, Publication & Ethical Review Board, with reference CHRPE/AP/172/19), and the Netherlands (Institutional Review Board of the AMC, University of Amsterdam, with reference NL32251.018.10). Participants gave informed consent to participate in the collection of the original data.

## RESULTS

The study involved a baseline sample of 2,183 participants of predominantly Akan ethnic origin (83.4%) comprising rural and urban Ghanaian dwellers who make up 29.7% and 28.2%, respectively. The remaining 42.1% of the study sample was made up of Ghanaian migrants living in Europe (Amsterdam) with a mean length of stay of 18.8 years. Out of the 2,183 participants, 102 did not have follow-up dietary data (loss to follow-up/non-response) comprising rural Ghana:10, urban Ghana:11 and Amsterdam:88 giving a dietary data follow-up response rate of 95.3%.

Table 1 shows the socio-demographic characteristics of the baseline study sample. Majority of the participants in rural Ghana were female (64.3%), married (58.9%), had only elementary education or never been to school (60.9%) and employed (84%) in the agricultural, forestry or fisheries sector (76.1%). The urban Ghana study site also had more female study participants (70.3%), more married people (63.7%) and (44.5%) had lower vocational/secondary schooling. Also, 84.7% of the participants living in urban Ghana were employed and mostly engaged in elementary occupations (42.3%) and the sales and services sector (22.8%). For Ghanaians living in Amsterdam, females as well constituted the greater proportion (60.7%) of the study participants. Further, 29.7% of the Amsterdam participants were divorced or separated and another 29.7% had never been married and 37.6% of them had education up to lower vocational/secondary school with 34.3% of them being employed and mostly (71.8%) engaged in elementary occupations.

**Table 1:**
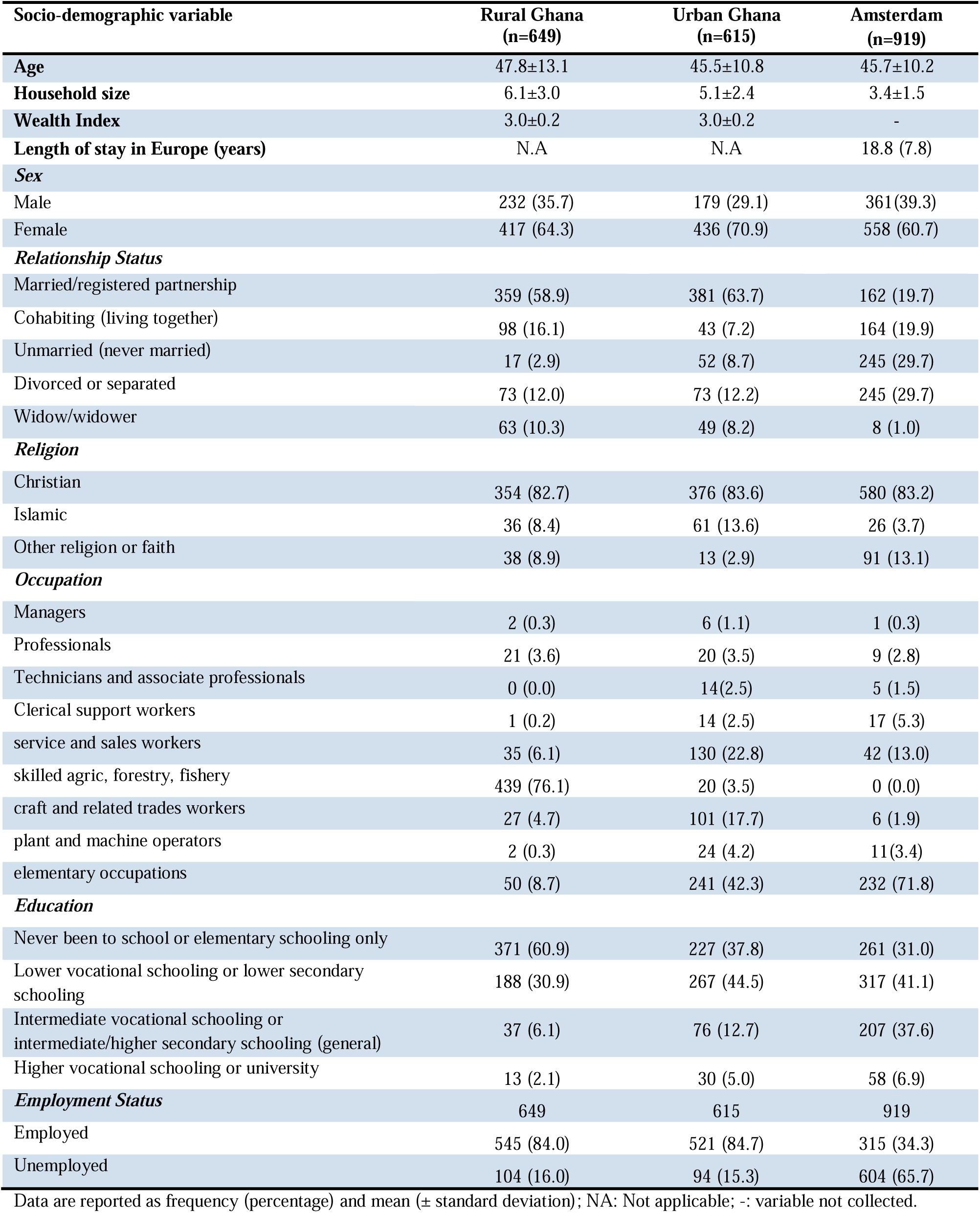
Socio-demographic Characteristics of study participants at Baseline.

### Change in daily caloric contribution of individual food groups

The daily caloric contribution of most of the individual food groups changed significantly between baseline and follow-up, with the exception of sweet spreads, dairy products, nuts and seeds, vegetables, alcoholic beverages and peanuts (Table 2). Individual food groups that recorded the highest significantly increased daily caloric contribution were fermented maize products (17.5%), whole grains and cereals (0.9%) and eggs (0.3%). On the other hand, coffee and tea, and refined cereals recorded the highest significantly decreased daily caloric contributions of −7.6% and −3.2% respectively.

**Table 2:**
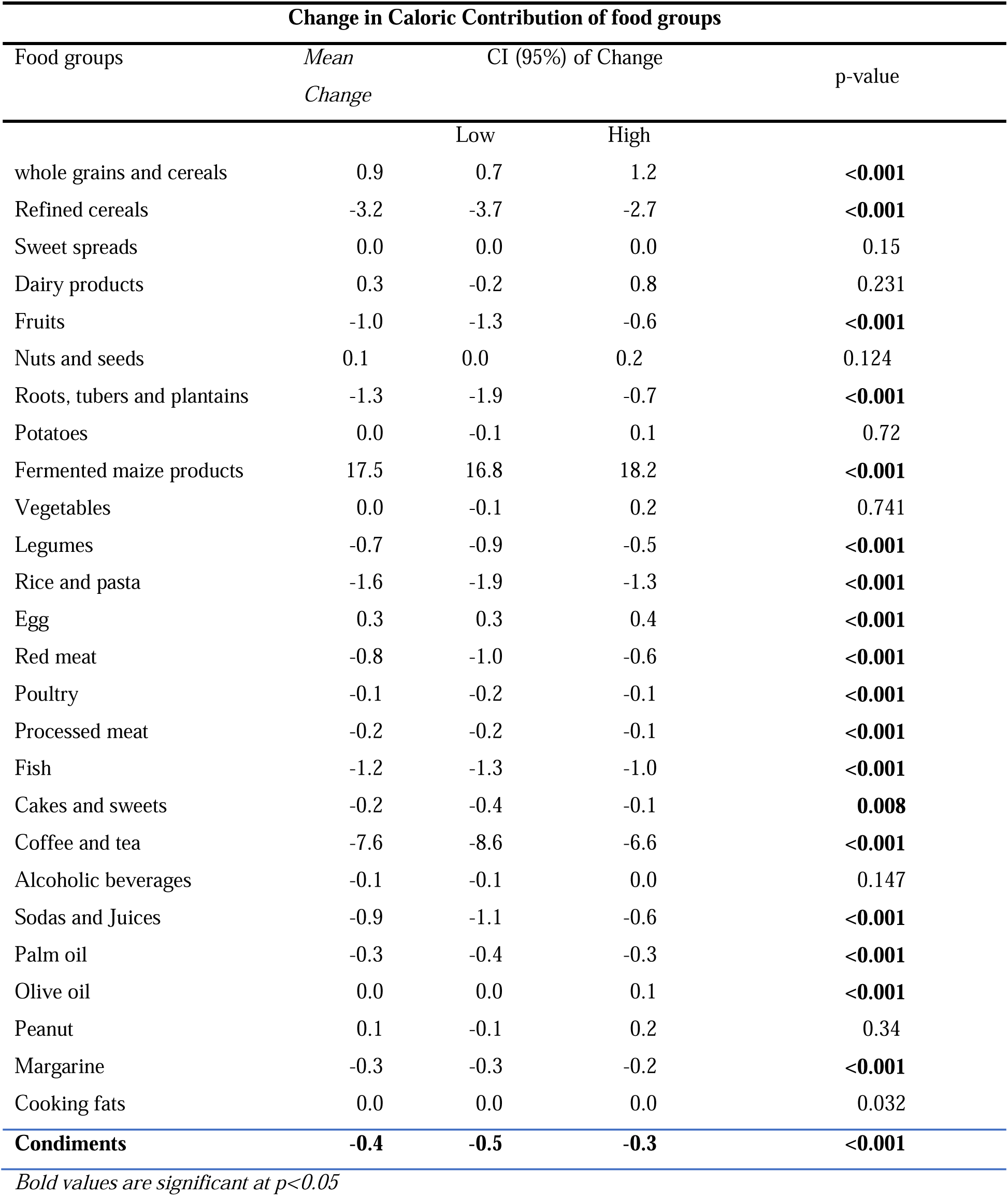
Change in daily caloric contribution of individual food groups.

Based on study site, the daily caloric contribution of the individual food groups with the highest changes were; whole grains and cereals, which increased by 1.1% in rural Ghana, 1.8% in urban Ghana and decreased by 0.9% in Amsterdam (Figure 1). Also, the daily caloric contribution of fermented maize products increased across all sites by 24.0%, 18.2% and 4.0% among rural, urban and Ghanaians in Amsterdam respectively. In contrast, the daily caloric contribution of coffee and tea decreased across all the three sites (Rural Ghana: −6.8%, Urban Ghana: −7.3% and Amsterdam: −9.8%).

**Figure 1:**
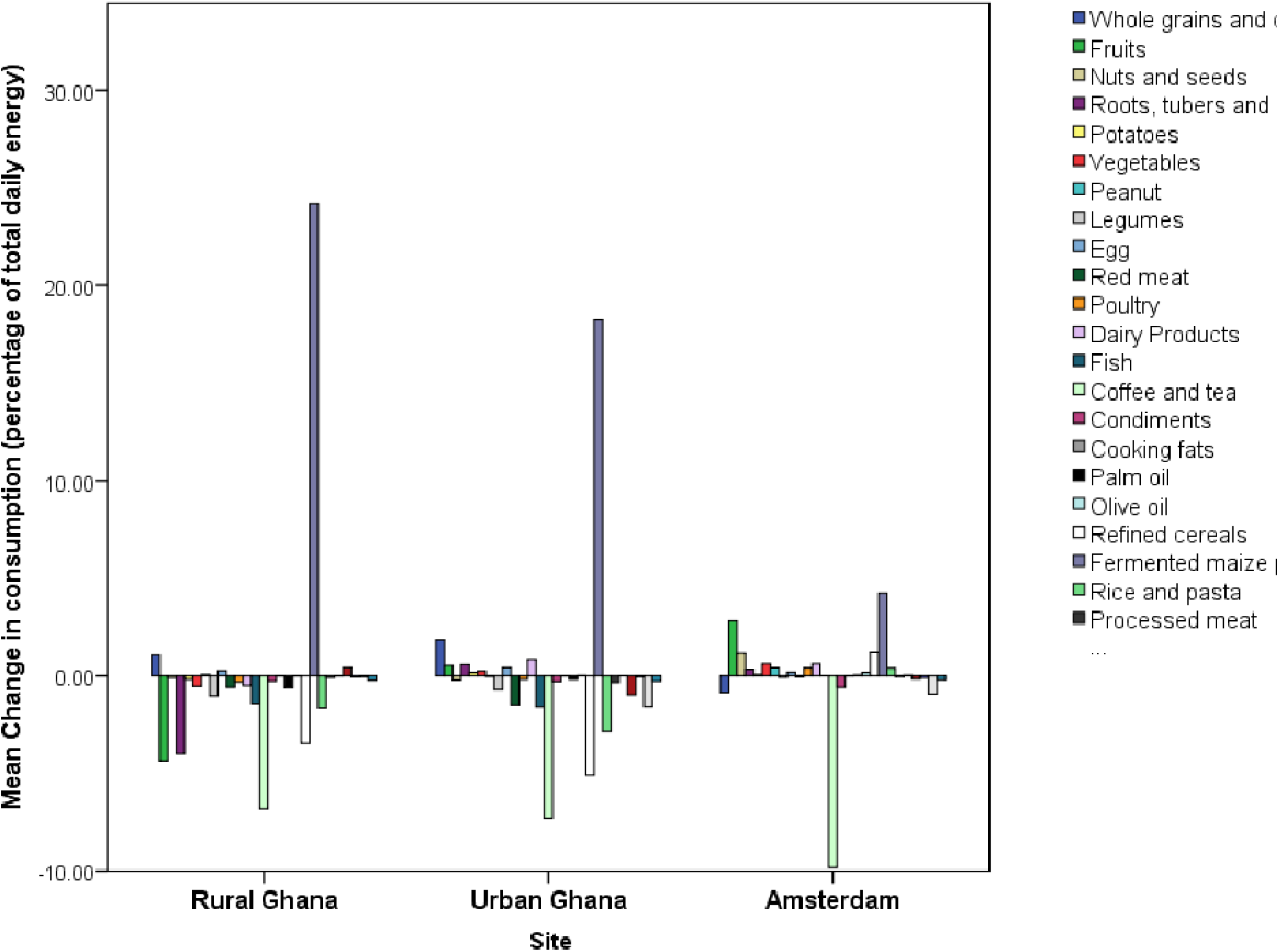
Mean change in consumption of food groups as a percentage of total daily energy according to study site.

### Changes in daily consumption of unprocessed, processed, and ultra-processed foods

The consumption of unprocessed/minimally processed foods among Ghanaians living in rural Ghana showed a significant decrease (p<0.001) by −21.2% (Mean Diff: −323.73g/day 95% CI: −408.78 - −238.69) from baseline intake and follow-up (Table 3). Conversely, for urban dwelling Ghanaians and Ghanaian migrants living in Amsterdam, the mean daily consumption of unprocessed/minimally processed foods increased significantly by 12.5% (Mean Diff::129.99g/day; 95%CI: 76.62 – 183.37, p<0.001) and 140.9% (Mean Diff: 1184.39g/day; 95%CI: 1079.37 −1289.41, p<0.001) respectively.

**Table 3.**
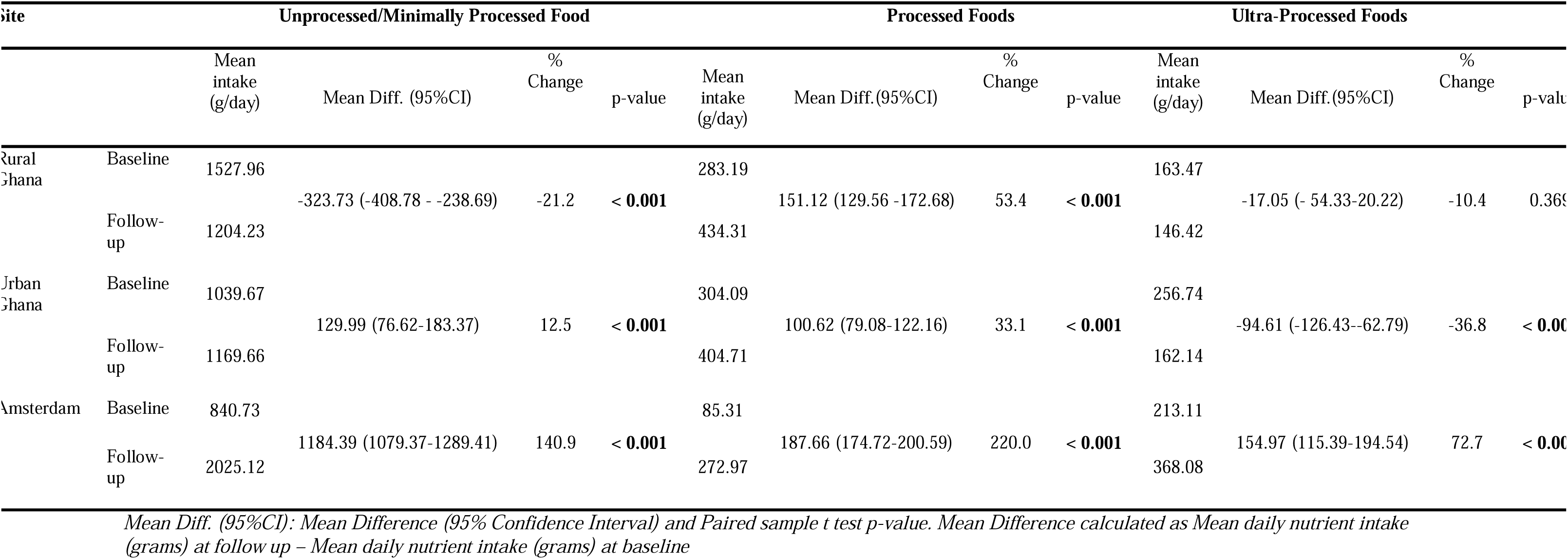
Changes over time in mean total daily intake (grams) of different levels of processed foods according to study site.

For processed foods, participants from rural Ghana increased their consumption by 53.4% (Mean Diff: 151.12g/day, 95%CI: 129.56 −172.68, p<0.001). Similarly, increased consumption of processed foods was observed among participants from urban Ghana (33.1% increase, Mean Diff: 100.62g/day, 95%CI: 79.08-122.16, p<0.001) and migrants living in Amsterdam (220.0% increase, Mean Diff: 187.66g/day, 95%CI: 174.72 −200.59, p<0.001).

The daily intake of ultra-processed foods on the other hand remained unchanged over the period for participants in rural Ghana. Whereas reduced intake of ultra-processed foods was observed among participants from urban Ghana (36.8% decrease, Mean Diff: −94.61g/day, 95%CI: −126.43 - −62.79, p<0.001), significantly increased consumption was recorded among migrant participants living in Amsterdam (72.7% increase, Mean Diff: 154.97g/day, 95%CI: 115.39 - 194.54, p<0.001).

### Changes in caloric contribution of unprocessed, processed and ultra-processed foods

After 6 years (±0.6) of follow-up, the estimated percentage of total daily energy from the consumption of unprocessed/minimally processed foods by participants in rural Ghana significantly decreased from 56.8% to 44.9% (% Change: −11.9%, 95%CI: −14.7 - −9.1, p<0.001) whereas the percentage of total daily energy from processed foods significantly increased from 20.8% to 39.5% (% change: 18.8%, 95%CI: 17.4 −20.1, p<0.001). However, the percentage of total energy from the consumption of ultra-processed foods did not significantly change (% Change: −0.6%, 95%CI: −1.4 - 0.2, p=0.136) among the rural dwellers in Ghana.

Similarly, the estimated percentage of total daily energy from the intake of unprocessed/minimally processed foods among the Urban Ghana cohort significantly decreased from 53.9% to 41.9% (% change: −12.0%, 95%CI: −14.2–– −9.7, p<0.001) whiles that of processed foods increased significantly from 23.6% to 33.5% (% change: 9.9%, 95%CI: 8.7 – 11.2, p<0.001). There was a significant decrease in the percentage of total daily energy from ultra-processed foods (% change: −2.0%, 95%CI: −3.0 – −1.1, p< 0.001).

For Ghanaian migrants in Amsterdam, the proportions of total daily energy from unprocessed/minimally processed foods and ultra-processed foods did not vary significantly between baseline and follow-up. On the other hand, a significant increase from 7.2% to 12.4% (% change from baseline intake: 5.3%, 95%CI: 44.3 – 66.3, p<0.001) was observed in the percentage of total daily energy from the consumption of processed foods among the Amsterdam group (Table 4).

**Table 4:**
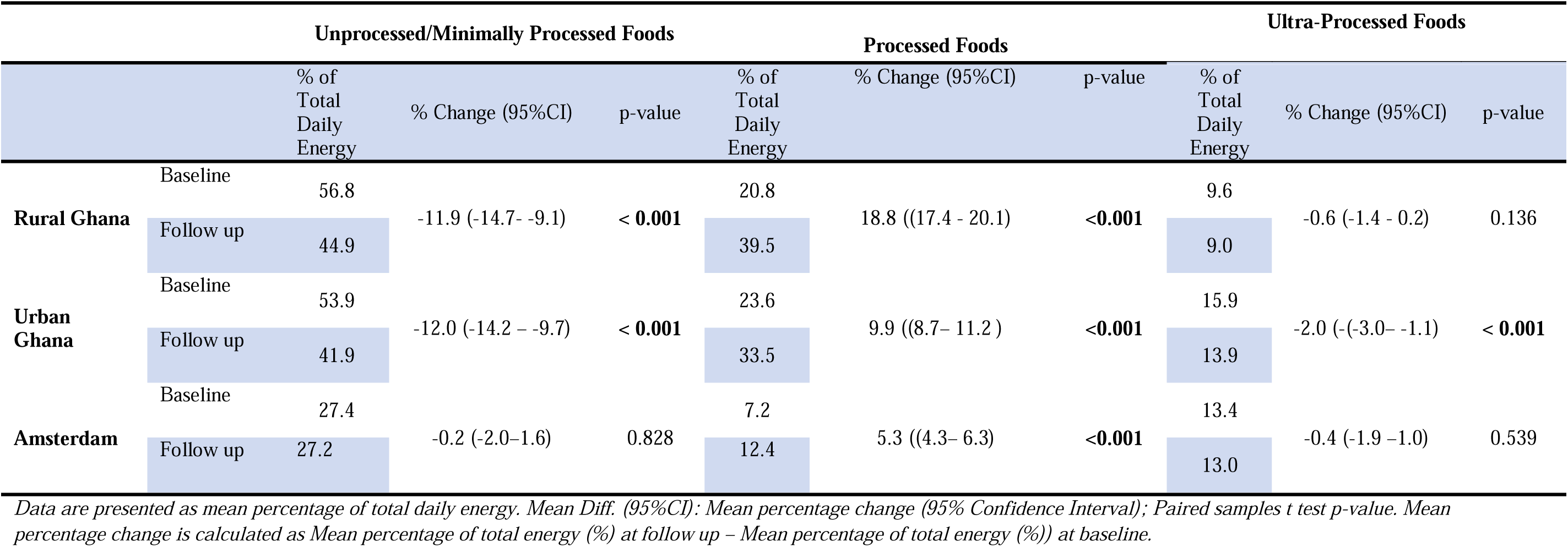
Changes in percentage of total daily energy intake from the different levels of processed foods among different Ghanaian populations.

An analyses of the socio-demographic drivers of the changes in the daily caloric contribution of the various levels of processed food shows that, for unprocessed/minimally processed foods, the decrease in caloric contribution occurred across sexes, educational level, relationship and employment status with the exception of unmarried people (p=0.287) and tertiary level participants (p=0.289) in Rural Ghana. In Urban Ghana, the decreased caloric contribution of unprocessed/minimally processed foods occurred across all socio-demographic groups while in Amsterdam, no significant change in caloric contribution was recorded across all groups with the exception of participants with secondary school/intermediate level of education whose caloric contribution from unprocessed foods increased by 3.2% (Supplementary Table S2). The caloric contribution of processed foods increased across all study sites and irrespective of socio-demographic status with the exception of Amsterdam where the widowed (p=0.287) and university level participants (p=0.372) had unchanged daily caloric contribution of processed foods between baseline and follow-up (Supplementary Table S3).

For ultra-processed foods, their caloric contribution was unchanged in Amsterdam and Rural Ghana across all socio-demographic groups whereas in Urban Ghana, the decrease in caloric contribution was observed only among males (−3.4%), married people (−2.9%) and people with secondary school educational level (−3.7%) (Supplementary Table S4).

## DISCUSSION

The findings from this study show a decline in the proportion of energy consumed from unprocessed/minimally processed food in both rural and urban Ghana, and no changes among migrant Ghanaians. A simultaneous increase in processed food consumption and in the contribution of processed foods to total energy intake was observed in each population group, confirming a common trend toward greater dependence on processed foods. The percentage of energy from ultra-processed foods among rural and migrant Ghanaians did not change, whereas energy contribution from ultra-processed food declined for urban Ghanaians.

The findings revealed that the quantities of intake of unprocessed/minimally processed foods significantly decreased among individuals living in rural Ghana resulting in a decrease in the percentage of energy from these foods. In contrast, there was a significant increase in the quantity of unprocessed foods consumed among Ghanaian urban dwellers and those living in Amsterdam, Netherlands. However, this did not lead to an increase in the percentage of energy but rather a decline among urban dwellers and unchanged for Ghanaian migrants in Amsterdam. This decrease in percentage of energy from unprocessed/minimally processed foods among rural and urban Ghanaians underscores the existence of the nutrition transition, which is characterized by the progressive shift from traditional diets mostly composed of unprocessed foods rich in fibre and low in fat and sugar to a diet made of processed and ultra-processed foods that are rich in animal products, refined grains, fats, salt and sugar but low in fibre [34]. For Ghanaian migrants in Amsterdam, the unchanged percentage of energy from unprocessed/minimally processed food in spite of their increased consumption could be a result of the increase in the overall caloric intake. It also seems possible that the unprocessed/minimally processed foods consumed by migrants may be in lower-energy form thus resulting in the unchanged percentage of energy from these foods.. According to Galbete et al. (2017), migration can also lead to dietary adaptations including the substitution of traditional foods with more energy-dense options in the host country [30] hence Ghanaian migrants may be replacing traditional staples, which are lower-energy unprocessed/minimally processed foods with higher energy unprocessed foods such as nuts and oily seeds resulting in the unchanged percentage energy from these foods.

A study by Cattafesta et al. (2020)[35] in Brazil among rural farmers ranked ultra-processed foods as one of the largest contributor to total calories (17.7%), being second to unprocessed/minimally processed foods (64.7%). While this study does not explicitly indicate a reduction in the consumption of unprocessed/minimally processed foods over time, it demonstrates that rural diets, which hitherto were largely made of unprocessed/minimally processed foods, now feature a significant proportion of processed foods. This position is buttressed by another recent evidence from a synthesis review of food systems transformations conducted by Baker et al. (2020)[36] which points to the issue of home-grown foods and unprocessed foods giving way to processed foods in rural areas in Africa and Asia due to increasing exposure to supermarkets and packaged food.

Our study also found that the daily consumption levels of processed foods has increased across all the different Ghanaian populations translating into an increased energy percentage contribution of processed foods to total energy of 19% among rural dwellers, 10% among urban dwellers and 5% among migrant Ghanaians in Amsterdam. Our findings of the increased daily consumption of processed foods among the different Ghanaian populations is largely supported by previous findings that there is a rise in the consumption of processed foods across West African countries [37].

The notable differences in change of consumption of unprocessed and processed foods across geographical locations likely reflects the large role of contextual environmental factors in shaping what people eat. Previous studies have shown that there is robust marketing and increased access to processed and foods [38–40] in especially urban areas leading to an over consumption of energy from these foods at the expense of unprocessed/minimally processed freshly prepared meals [41]. Also, it has been noted in Ghana that, processed foods account for the largest share of all foods in the urban food environment where 80% of retail outlets sell processed foods. Processed foods also constitute the most dominant food category in retail outlets making it highly accessible in equal measure as ultra-processed foods [42]. For the increase in the intake of processed foods and the rise in percentage of calories from these foods in rural areas in Ghana, this may be as a result of factors such as increased incomes from non-farm activities as well as increased mechanization of farm production and the situation where more women are now working outside their homes as suggested by the International Food Policy Research Institute (IFPRI) [43]. Further, in rural areas in Ghana, processed staple foods such as processed maize meals, gari (grated cassava), smoked fish among others may also be more cheaper than imported packaged snacks and frozen meals [44] making people to rely more on the traditionally processed foods instead. Previous studies have also shown that migrants in Europe of Ghanaian origin mostly retain their traditional food choices [45], which mostly involves the use of processed ingredients such as dried fish, gari, etc. They may however be replacing these ingredients with more processed options such as tomato paste and canned fish among others due to availability.

With respect to ultra-processed foods, whereas consumption levels remained unchanged for the rural Ghana population, there was decline in consumption and percentage contribution to total energy among the urban Ghanaian population. In contrast to these findings where the percentage of energy from ultra-processed foods among urban dwellers in Ghana declined from 15.9% to 13.9%, an almost concurrent study conducted between 2008–2009 and 2017–2018 in Brazil showed an increase in the percentage energy from ultra-processed foods in urban areas from 19.94% to 20.55% [46]. The differences in the dietary recall periods and characteristics of the study subjects may have partly accounted for the different patterns of caloric intake from ultra-processed foods in the two studies. Most of the previous studies that have observed increasing consumption levels of ultra-processed foods in urban areas have cited various reasons including aggressive marketing [38], increased physical availability [39] and affordability [40].

There is a global rise in the consumption of ultra-processed foods as they already constitute the highest contributors to daily calories in high-income countries such as the United Kingdom [47], the United States [48], Canada [49] and Australia [50]. Similarly, our study points to a rising daily intake of ultra-processed food with levels shifting to the higher consumption brackets among Ghanaians living in the Netherlands although we noted that the proportion of total energy contributed by ultra-processed foods among this population living in a high-income country has not significantly increased over the period. We hypothesize that the apparent stagnation in the percentage of calories from ultra-processed foods among this group of migrants may be related to a saturation effect where the consumption of ultra-processed foods has stabilized after an initial high increase when they first became exposed to the Dutch food environment. Otherwise, this could be due to a slower nutrition transition inspired by a lower level of acculturation [45]..

This study provides valuable insights into the longitudinal changes in processed food intake among populations experiencing rapid nutrition transitions. The strengths include its prospective design, and stratification of the study population according rural, urban and migrants, which helped to assess the environmental influence on food and energy intake. Another strength is that there was standardization of the data collection process across study sites where the same methods were used in Amsterdam, rural and urban Ghana. We also used individual-level dietary survey data, which estimated actual food consumption unlike in other studies that used food availability and statistics of production and trade, which may not provide a completely accurate reflection of dietary consumption. The use of the NOVA food classification system is also a plus since it has been recognized by UN agencies as a relevant approach for linking dietary intake, obesity and Non-Communicable Diseases (NCDs) [41,51,52].

This study, however, is not without limitations. First, as may be the case for most dietary studies, the limitation of self-reporting and recall bias leading to misreporting cannot be ruled out in this study. It’s effect however on the study outcomes may be minimal due to the standardization of the data collection process across the sites. Secondly, the original dietary survey was not designed specifically to categorize foods according to characteristics of industrial processing, and so some misclassification of foods at the individual level cannot be excluded given that we applied the classification at the level of food groups rather than individual food items, which could lead to some overestimation or underestimation of the energy contribution from the various food groups. However, we minimized classification errors by setting standardized, objective and clear criteria, and a conservative approach (assigning lower level of processing) was used in case of uncertainty.

## Conclusion

This study concludes that dietary changes are occurring within population groups in Ghana, as part of a broader nutrition transition. Our findings indicate that the rate of increase in processed foods differs by geographical location, suggesting an important influence of contextual factors. This underscores the need for targeted strategies to curb the rising intake of these potentially harmful foods. Further research is necessary to identify the specific contextual drivers of this increase, particularly among Ghanaian migrants in Europe, to inform the development of tailored interventions.

## Data Availability

Data described in the manuscript and code book will be made available upon request pending application and approval. Requests should be directed to Dr Erik Beune or Prof Charles Agyemang.

## Acknowledgments

We thank the research assistants, interviewers and other staff of the three research locations who have taken part in gathering the data and, most of all, the Ghanaian volunteers participating in this project. We gratefully acknowledge Amsterdam UMC, AMC-Biobank for their support in biobank management and high-quality storage of collected samples.

