## Supplementary Table S1 for "Longitudinal changes in processed foods intake and their daily caloric contribution among Ghanaian populations living in Ghana and Europe: Findings from the prospective RODAM Cohort study"

Supplementary Table S1: Description of food groups and NOVA classification

| **Food group** | **Description** | **Processing Levels according to NOVA Classification** |
| --- | --- | --- |
| Whole grains and cereals | Whole grain bread, wholegrain crispbread, muesli cereals, and other grains (millet, couscous, polenta, spelt, and barley) | G1: Unprocessed/minimally processed |
| Fruits | Orange, mandarin, kiwi, watermelon, mango, cantaloupe, pawpaw, pineapple, banana, plum, peach, apricot, nectarine, flat peach, apple, pear, strawberries, cherries, berries, grapes, and stewed fruit | G1: Unprocessed/minimally processed |
| Peanut | Peanut, peanut butter | G1: Unprocessed/minimally processed |
| Nuts and seeds | Dried fruit, nuts, and seeds | G1: Unprocessed/minimally processed |
| Roots, tubers and plantain | Plantain, cassava, yam, and fufu | G1: Unprocessed/minimally processed |
| Potatoes | Potatoes, pan fried potatoes, French fries, and sweet potatoes | G1: Unprocessed/minimally processed |
| vegetables | Green leaves, spinach, chard, lettuce, endive, chicory, Chinese and white cabbage, tomatoes, peppers, carrots, cucumber, eggplant, beans (green beans), onions and garlic | G1: Unprocessed/minimally processed |
| Legumes | Groundnut soup, legumes, lentil-pea and bean soup | G1: Unprocessed/minimally processed |
| Egg | Egg | G1: Unprocessed/minimally processed |
| Red meat | Beef, goat, pork, game, liver, and giblets | G1: Unprocessed/minimally processed |
| Poultry | Poultry | G1: Unprocessed/minimally processed |
| Fish | Fatty fish, lean fish, fish preparations and shellfish | G1: Unprocessed/minimally processed |
| Coffee and tea | Regular coffee, decaffeinated coffee, black and green tea, and fruit and herbal tea | G1: Unprocessed/minimally processed |
| Olive oil | Olive oil | G2: Processed culinary ingredients |
| Palm oil | Palm oil | G2: Processed culinary ingredients |
| Cooking fats | Cooking fats (e.g. animal fats like lard or speck) | G2: Processed culinary ingredients |
| condiments | Ketchup, mayonnaise, crème fraiche, salad cream, sour cream, remoulade, and sauces | G2: Processed culinary ingredients |
| Fermented maize products | Banku and kenkey | G3: Processed foods |
| Refined cereals | White wheat bread, white crispbread, hot cereals, and porridge | G3: Processed foods |
| Rice and pasta | Rice, pasta, noodles, and macaroni | G3: Processed foods |
| Processed meat | Meatballs, fried sausage, boiled sausage, dry and cured meat, salami, jagdwurst, bologna, mortadella, ham corned beef, liverwurst, and liver pâté | G3: Processed foods |
| Cakes and sweets | Tart, pie, yeast cake, pastry, sponge cake, cream pie, cheesecake, cookies, chocolate, sweets, candy, and toffee | G4: Ultra-processed foods |
| Sweet spreads | Marmalade, jam, jelly, and honey | G4: Ultra-processed foods |
| Dairy products | Cocoa milk drink, fruit milk drink, plain yoghurt, buttermilk, flavoured yoghurt, soft cheese, semi-soft/firm cheese, sour milk, quark, mozzarella, mascarpone, feta cheese, butter, whipped cream | G4: Ultra-processed foods |
| Alcoholic beverages | Regular beer, wine, liquors, and spirits | G4: Ultra-processed foods |
| Sodas and juices | Non-alcoholic beer, sodas and minerals, light and soft drinks, fruit juices, fruit nectars, vegetable juices | G4: Ultra-processed foods |
| Vegetable soups, stews, sauces | Palmnut soup, nkontomire stew, okro stew, tomato sauce and stew, vegetable soup | Mixed classification |
