## Supplementary Table S2 for "Longitudinal changes in processed foods intake and their daily caloric contribution among Ghanaian populations living in Ghana and Europe: Findings from the prospective RODAM Cohort study"

Supplementary Table S2: Changes in percentage of total daily energy intake of Unprocessed/Minimally processed foods according to study site and socio-demographic and factors

| Socio-demographic | RURAL GHANA | | URBAN GHANA | | | AMSTERDAM | | |
| --- | --- | --- | --- | --- | --- | --- | --- | --- |
|  | % Change (95%CI) | p-value | | % Change (95%CI) | p-value | | % Change (95%CI) | p-value |
| Overall | -11.9 (-14.7- - 9.1) | **< 0.001** | | -12.0 (-14.2 - -9.7) | **<0.001** | | -0.2 (-2.0 – 1.6) | 0.828 |
| Male | -11.7(-16.0 – 7.5) | **<0.001** | | -6.1 (-9.8 - -2.4) | **0.001** | | -0.3 (-3.3 – 2.7) | 0.845 |
| Female | -12.0 (-15.6 - -8.4) | **<0.001** | | -14.4 (-17.2 - -11.6) | **<0.001** | | -0.1 (-2.3 – 2.1) | 0.906 |
| Married | -12.0 (-15.7 - -8.2) | **<0.001** | | -10.4 (-12.9 - -7.9) | **<0.001** | | 0.3 (-3.7 – 4.3) | 0.893 |
| Cohabiting | -8.4 (-14.0 - -2.8) | **0.004** | | -18.2 (-27.6 - - 8.7) | **<0.001** | | 1.0 (-2.4 -4.5) | 0.551 |
| Unmarried | -0.3 (-15.1 – 14.5) | 0.966 | | -17.8 (-26.7 - -9.0) | **<0.001** | | -0.6 (-4.7 – 3.6) | 0.782 |
| Divorced | -8.2 (-16.3 - -0.1) | **0.048** | | -11.5 (-17.6 - -5.3) | **<0.001** | | -0.7 (-4.3 – 2.8) | 0.681 |
| Widow/widower | -14.9 (-27.4 - -2.4) | **0.020** | | -20.7 (-34.5 - - 6.8) | **0.004** | | -3.6 (-29.8 – 22.6) | 0.693 |
| Never /elementary | -11.4 (-15.3 - -7.5) | **<0.001** | | -18.6 (-23.0 - - 14.3) | **<0.001** | | 0.4 (-2.8 – 3.7) | 0.787 |
| Lower voc/secondary | -9.4 (-14.3 - -4.4) | **<0.001** | | -8.1 (-10.9 - - 5.2) | **<0.001** | | -1.5 (-4.6 – 1.6) | 0.346 |
| Intermediate /second | -15.1 (-23.9 - - 6.3) | **0.001** | | -11.4 (-16.6 - -6.3) | **<0.001** | | 3.2 (0.2 – 6.1) | **0.039** |
| university | -8.8 (-26.0 – 8.4) | 0.289 | | -7.0 (-19.8 – 5.8) | 0.273 | | -4.7 (-12.4 – 2.9) | 0.212 |
| Employed | -11.0 (-14.1 - -7.8) | **<0.001** | | -12.9 (-15.6 - -10.1) | **<0.001** | | -0.4 (-3.0 – 2.2) | 0.768 |
| Unemployed | -10.3 (-18.5 - -2.2) | 0.014 | | -12.8 (-17.4 - - 8.2) | <0.001 | | 1.1 (-3.5 – 5.8) | 0.626 |

*% Change. (95%CI): Mean Percentage change (95% Confidence Interval) calculated as mean caloric contribution at follow-up minus mean caloric contribution at baseline. P-value: Paired sample t test p-value with significant values highlighted*
