## Supplementary Table S3 for "Longitudinal changes in processed foods intake and their daily caloric contribution among Ghanaian populations living in Ghana and Europe: Findings from the prospective RODAM Cohort study"

Supplementary Table S3: Changes in percentage of total daily energy intake of processed foods according to study site and socio-demographic factors

| Socio-demographic | RURAL GHANA | | URBAN GHANA | | | | | AMSTERDAM | |
| --- | --- | --- | --- | --- | --- | --- | --- | --- | --- |
|  | % Change (95%CI) | p-value | | % Change (95%CI) | | p-value | % Change (95%CI) | | p-value |
| Overall | 18.8 (17.4 – 20.1) | **<0.001** | | | 9.9 (8.7 – 11.2) | **<0.001** | 5.3 (4.3 – 6.3) | | **<0.001** |
| Male | 19.4 (17.1 – 21.6) | **<0.001** | | 10.9 (8.6 – 13.3) | | **<0.001** | 5.7 (4.0 – 7.4) | | **<0.001** |
| Female | 18.4 (16.7 – 20.1) | **<0.001** | | 9.5 (8.0 – 11.0) | | **<0.001** | 5.0 (3.8 – 6.2) | | **<0.001** |
| Married | 18.3 (16.4 – 20.1) | **<0.001** | | 10.5 (8.8 – 12.1) | | **<0.001** | 5.2 (3.1 – 7.3) | | **<0.001** |
| Cohabiting | 20.7 (17.2 – 24.2) | **<0.001** | | 9.1 (3.4 – 14.8) | | **0.003** | 6.8 (4.6 – 9.0) | | **<0.001** |
| Unmarried | 18.4 (9.7 – 27.1) | **<0.001** | | 8.0 (4.7 – 11.3 ) | | **<0.001** | 4.6 (3.0 – 6.2) | | **<0.001** |
| Divorced | 19.3 (15.3 – 23.3) | **<0.001** | | 9.3 (5.4 – 13.3) | | **<0.001** | 5.1 (2.8 – 7.4) | | **<0.001** |
| Widow/widower | 20.3 (16.0 – 24.7) | **<0.001** | | 9.5 (5.0 – 14.0) | | **<0.001** | 5.2 (-7.6 – 17.9) | | 0.287 |
| Never /elementary | 18.8 (17.0 – 20.6) | **<0.001** | | 9.8 (7.7 – 11.9) | | **<0.001** | 5.9 (4.0 – 7.7) | | **<0.001** |
| Lower voc/secondary | 19.2 (16.6 – 21.8) | **<0.001** | | 10.5 (8.6 – 12.5) | | **<0.001** | 5.2 (3.6 – 6.8) | | **<0.001** |
| Intermediate /second | 21.1 (16.2 – 26.1) | **<0.001** | | 8.9 (5.3 – 12.5) | | **<0.001** | 6.2 (4.2 – 8.2) | | **<0.001** |
| university | 16.8 (6.1 – 27.6) | **0.005** | | 8.0 (4.0 – 12.0) | | **<0.001** | 1.4 (-1.8 – 4.6) | | 0.372 |
| Employed | 19.4 (17.9 – 20.8) | **<0.001** | | 10.2 (8.5 – 11.8) | | **<0.001** | 6.0 (4.6 – 7.3) | | **<0.001** |
| Unemployed | 15.3 (9.8 – 20.8) | <0.001 | | 9.8 (7.5 – 12.1) | | <0.001 | 4.1 (1.7 – 6.5) | | 0.002 |

*% Change. (95%CI): Mean Percentage change (95% Confidence Interval) calculated as mean caloric contribution at follow-up minus mean caloric contribution at baseline. P-value: Paired sample t test p-value with significant values highlighted*
