## Supplementary Table S4 for "Longitudinal changes in processed foods intake and their daily caloric contribution among Ghanaian populations living in Ghana and Europe: Findings from the prospective RODAM Cohort study"

Supplementary Table S4: Changes in percentage of total daily energy intake of Ultra-processed foods according to study site and socio-demographic factors

| Socio-demographic | RURAL GHANA | | URBAN GHANA | | | AMSTERDAM | | |
| --- | --- | --- | --- | --- | --- | --- | --- | --- |
|  | % Change (95%CI) | p-value | | % Change (95%CI) | p-value | % Change (95%CI) | | p-value |
| Overall | -0.6 (-1.4 – 0.2) | 0.136 | | -2.0 (-3.0 - -1.1) | **<0.001** | -0.4 (-1.9 – 1.0) | | 0.539 |
| Male | -0.3 (-1.7 – 1.2) | 0.727 | | -3.4 (-5.4 - -1.5) | **<0.001** | -1.1 (-3.4 – 1.2) | | 0.333 |
| Female | -0.8 (-1.7 – 0.1) | 0.098 | | -1.5 (-2.6 - - 0.3) | **0.013** | -0.1 (-1.9 – 1.8) | | 0.980 |
| Married | 0.4 (-0.6 – 1.5) | 0.411 | | -2.9 (-4.2 - - 1.7) | **<0.001** | -1.0 (-4.4 – 2.3) | | 0.536 |
| Cohabiting | -0.8 (-2.8 – 1.1) | 0.408 | | 1.9 (-1.2 – 5.0) | 0.215 | -1.0 (-4.0 – 2.0) | | 0.495 |
| Unmarried | -7.1 (-13.3 - -0.9) | **0.027** | | -1.2 (-4.8 – 2.4) | 0.518 | 2.5 (-0.9 – 5.9) | | 0.144 |
| Divorced | -2.2 (-4.5 - - 0.2) | 0.067 | | -0.4 (-2.9 – 2.1) | 0.734 | -0.5 (-2.9 – 1.8) | | 0.641 |
| Widow/widower | -1.5 (-4.0 – 1.0) | 0.241 | | -2.1 (-5.3 – 1.1) | 0.198 | -17.3 (-37.1 – 2.5) | | 0.069 |
| Never /elementary | -0.2 (-1.2 – 0.8) | 0.753 | | -0.2 (-1.8 – 1.3) | 0.755 | 0.1 (-2.6 – 2.7) | 0.947 | |
| Lower voc/secondary | -1.2 (-2.7 - - 0.3) | 0.105 | | -3.7 (-5.1 - - 2.2) | **<0.001** | -1.0 (-3.6 – 1.5) | | 0.430 |
| Intermediate /second | -0.9 (-4.5 – 2.8) | 0.627 | | -2.0 (-5.1 – 1.2) | 0.217 | -0.4 (-3.0 – 2.2) | | 0.755 |
| university | 1.6 (-7.0 – 10.1) | 0.694 | | -1.3 (-5.6 – 3.0) | 0.553 | 0.1 (-5.5 – 5.6) | | 0.977 |
| Employed | -0.5 (-1.3 – 0.4) | 0.258 | | -2.1 (-3.3 – 0.9) | **0.001** | -1.6 (-3.7 – 0.5) | | 0.139 |
| Unemployed | -1.0 (-4.3 – 2.3) | 0.535 | | -2.3 (-4.1 - - 0.5) | 0.012 | 0.5 (-3.1 – 4.1) | | 0.786 |

*% Change. (95%CI): Mean Percentage change (95% Confidence Interval) calculated as mean caloric contribution at follow-up minus mean caloric contribution at baseline. P-value: Paired sample t test p-value with significant values highlighted*
